# Can social prescribing reach patients most in need? Patterns of (in)equalities in referrals in a representative cohort of older adults in England

**DOI:** 10.1101/2024.03.26.24304891

**Authors:** Daisy Fancourt, Andrew Steptoe

**Affiliations:** Department of Behavioural Science and Health, University College London

## Abstract

**Importance:** Social prescribing (SP) is a mechanism of care referring people to non-clinical forms of support and services in local communities to improve health and wellbeing. But there is much contention over whether SP is in fact provided disproportionately more to individuals who are less disadvantaged. However, a comprehensive analysis of who is receiving SP from both medical and non-medical referral routes has never been undertaken.

**Objective:** To incorporate bespoke novel questions on SP into a nationally-representative cohort study to assess whether SP is truly reaching individuals most in need.

**Design:** We used data from Wave 10 (2021/23) of the English Longitudinal Study of Ageing (ELSA) involving richly-phenotyped adults aged 50+ living in England. We explored how SP was patterned according to social and health inequalities, all measured in Wave 9 (2018/19; prior to the reported engagement in SP). Multiple ordinal regression models were used to explore predictors of receiving a SP referral.

**Setting:** Community-dwelling older adults living in England

**Participants:** 7,283 adults aged 50+ answered the self-completion questionnaire at wave 10 so were included in analyses.

**Main outcomes and measures:** Participants were asked if they had received a referral to a wide range of community-based activities by a doctor, social worker or other health professional

**Results:** 495 adults (6.8%) reported receiving a SP referral, and 435 (88%) accepted. Age was a significant predictor of referrals (OR 1.02, CI 1.01-1.03), but being female was not (OR 1.05, CI 0.84-1.30). Receiving benefits increased referral odds two-fold (OR 2.03, CI 1.53-2.70) and being in the lowest wealth tertile (relative to middle tertile) by 55% (CI 1.13-2.13). Education and employment status did not predict referrals. Referrals were most common for individuals with depression (OR=1.60, CI 1.20-2.12), diabetes (OR=1.52, CI 1.11-2.10), chronic pain (OR=1.57, CI 1.22-2.02), multiple long-term conditions (4+ conditions, OR=2.8, CI 1.41-5.51), and who were physically inactive (OR=1.63, CI 1.10-2.41).

**Conclusions and relevance:** There is some initial evidence of SP referrals occurring amongst older adults in England, with high uptake amongst those referred. Promisingly, those with highest socio-economic need and most long-term health conditions particularly appear to be receiving support.

**Key points:** *Question:* Who is receiving referrals to social prescribing programmes?

*Findings:* In this analysis of a nationally representative cohort study of 7,283 richly-phenotyped adults aged 50+ living in England, SP was offered most to individuals experiencing socio-economic adversity with health needs including depression, diabetes, chronic pain, multiple long-term conditions, and those who were physically inactive.

*Meaning:* Promisingly, older adults with highest socio-economic need and most long-term health conditions particularly appear to be receiving support through social prescribing.

## Introduction

Social prescribing (SP) is a mechanism of care referring people to non-clinical forms of support and services in local communities to improve health and wellbeing ^1^. While initially traced back to initiatives developed in the UK, SP is now gaining global traction, with many countries implementing programmes to address social determinants of health, with the aim of reducing health inequalities ^2^. However, this claim has proved controversial, with concerns that SP cannot address upstream determinants of inequalities and could even be provided disproportionately more to individuals who are less disadvantaged ^3–5^.

While SP (as an individual-level intervention) cannot address upstream determinants of inequalities, it does broaden existing health service provision and could increase uptake of community activities amongst those least likely to engage, as well as supporting self-management of existing health conditions ^6^. The challenge is that few studies have analysed issues of uptake and equity ^7^.

In the UK, SP is included in the NHS Long Term Plan for “tackling health inequalities”, and its national roll-out as a programme across England provides a fertile opportunity to explore equity of access. However, it is a major challenge to assess who is receiving SP. Local evaluations can be too small in scale, while electronic patient records often contain inconsistent coding of SP referrals, poor recording of wider determinants of health, and no details of referrals from sources other than GPs, leading to conclusions that they cannot be used to assess equity of referrals ^8,9^. Thus, exploring SP referral patterns in wider data sources is crucial to understanding if SP is truly reaching individuals most in need.

In this study, we incorporated bespoke questions on SP into a nationally representative cohort study of richly-phenotyped adults aged 50+ living in England to identify patterns of SP referrals and their uptake.

## Methods

We used data from Waves 9 and 10 of the English Longitudinal Study of Ageing ^10^. Questions on SP in Wave 10 (2021/23) asked “In the past 2 years, has a doctor, social worker or other health professional referred you to take part in any of the following: (i) arts, crafts, music, reading groups, or social groups, (ii) gardening or nature activities, (iii) outdoor health or fitness activities/clubs, (iv) indoor exercise or other clubs/activities, (v) adult learning or skills development training, (vi) employment or benefit support, or (vii) other social, community or volunteering activity.” Responses were “No”, “Yes, but I did not accept”, “Yes, I attended just one session” or “Yes, I attended more than one session”. 7,283 adults aged 50+ answered the self-completion questionnaire at wave 10 so were included in analyses.

We explored how SP was patterned according to social and health inequalities, all measured in Wave 9 (2018/19; prior to the reported engagement in SP). Social inequalities included sex, age (years), marital status (married/cohabiting vs other), educational attainment (no qualifications/basic vocational qualifications vs GCE/O level vs A-level vs degree), retirement status, net non-pension wealth (tertiles), receipt of any state benefits, urban living (yes vs no), and area deprivation (from linking lower layer super output area data to Index of Multiple Deprivation from the 2021 census). Health inequalities included a diagnosed psychiatric condition, depressive symptoms (≥3 on the 8-item Center for Epidemiologic Studies Depression Scale; CES-D), diagnosed cardiovascular condition (hypertension, angina, heart attack, heart failure, heart murmur, abnormal heart rhythm, stroke or heart disease), diabetes, respiratory condition (lung disease or asthma), musculoskeletal condition (arthritis or osteoporosis), dementia or cancer, sedentary behaviour (any physical activity less than once a week), alcohol consumption (≥5 times a week), current smoking, and chronic pain.

Multiple ordinal regression models were used to explore predictors of receiving a SP referral. Multiple imputation by chained equations was used to provide 30 imputed datasets, incorporating all predictors and identified auxiliary variables relating to social behaviours. Main analyses binarized SP as receiving a SP referral (regardless of uptake) and sensitivity analyses instead explored uptake vs not/no referral. Model 1 was adjusted for age and sex, model 2 = model 1 + social inequalities, model 3 = model 1 + health inequalities, and model 4 = model 1 + social and health inequalities.

## Results

495 adults (6.8%) reported receiving a SP referral, and 435 (88%) accepted. Exercise classes were the most frequent intervention, followed by arts groups, nature-based activities, and adult learning. Of those referred to SP, 56% were female, average age 67.1 (SE 0.5 years) (see supplementary table 1 for full demographics). Results from the first model in which variables were included are reported below. Full tables of results are in supplementary table 2.

Age was a significant predictor of referrals (OR 1.02, CI 1.01-1.03), but being female was not (OR 1.05, CI 0.84-1.30). SP was offered most to individuals experiencing socio-economic adversity. Receiving benefits increased referral odds two-fold (OR 2.03, CI 1.53-2.70) and being in the lowest wealth tertile (relative to middle tertile) by 55% (CI 1.13-2.13). Education and employment status did not predict referrals.

Referrals were most common for individuals with depression (OR=1.60, CI 1.20-2.12), diabetes (OR=1.52, CI 1.11-2.10), chronic pain (OR=1.57, CI 1.22-2.02), and who were physically inactive (OR=1.63, CI 1.10-2.41). Diagnosis of psychiatric, cardiovascular, respiratory, and musculoskeletal conditions did not predict referrals individually, nor did cancer, smoking status or alcohol consumption. However, when substituting specific health conditions with a count variable of the number of multiple long-term conditions, those with more conditions were more likely to be referred (4+ conditions, OR=2.8, CI 1.41-5.51).

Results were all maintained in model 4, although physical inactivity became a less strong predictor (see Figure 1 & supplementary table 2). Sensitivity analyses focusing on uptake of SP (rather than just referral) produced very similar findings (see supplementary table 3).

**Figure 1:**
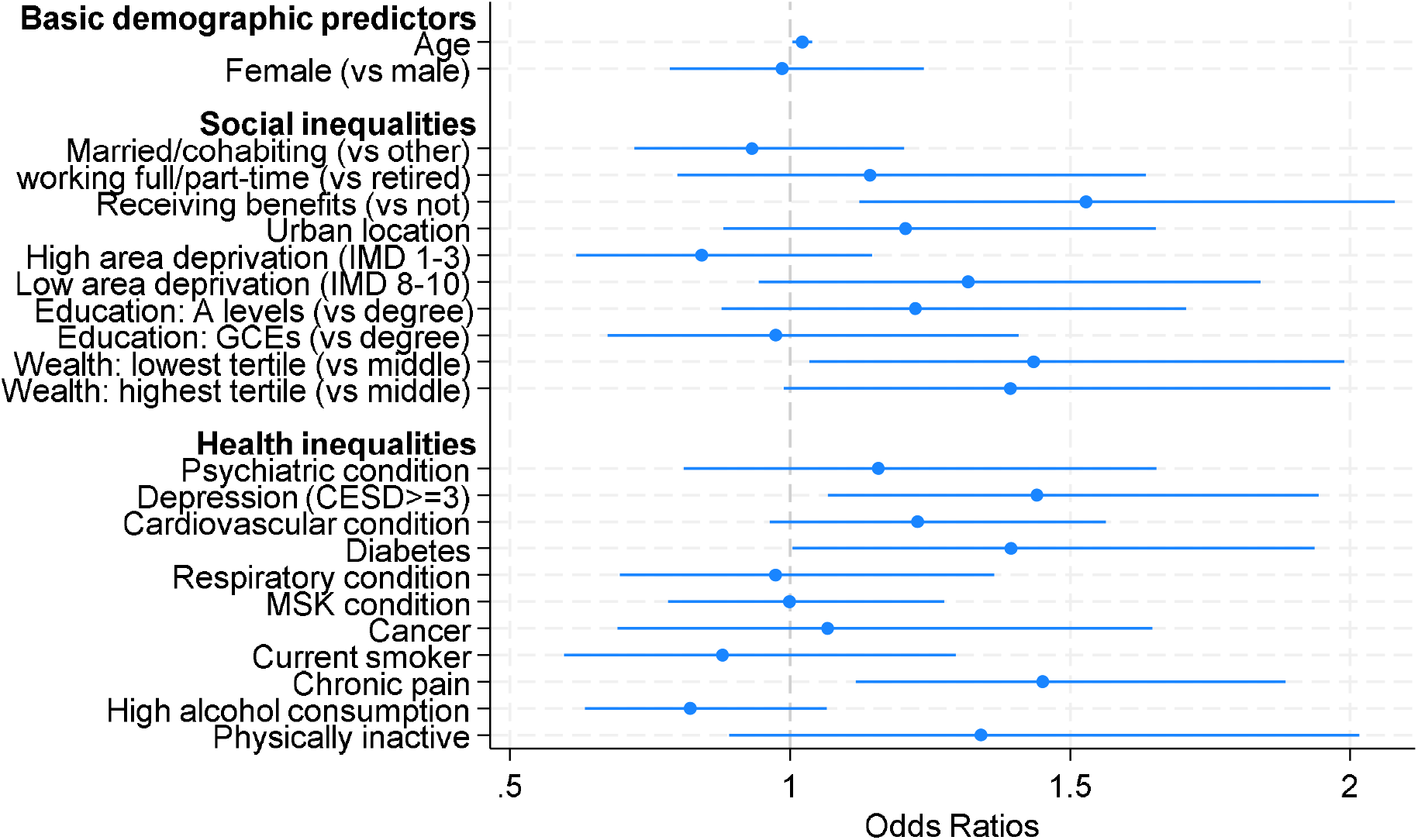
Predictors of SP referrals in fully-adjusted ordinal regression models (model 4)

## Discussion

There is some initial evidence of SP referrals occurring amongst older adults in England, with high uptake amongst those referred.

Promisingly, those with highest socio-economic need and most long-term health conditions particularly appear to be receiving support. Prior research has shown these groups as the least likely to engage in community-based activities like those offered by social prescribing, as well as the most likely to experience poor long-term health ^11^. This suggests that SP does have the potential for reaching individuals who are more disadvantaged. However, further research is needed to explore how initial uptake of referrals translates to subsequent engagement patterns and, in particular, whether inequalities shape participants’ capacity to benefit from the interventions provided ^12^.

SP relies on participant self-report of referrals, which is subject to bias. However, overall, these data from a representative cohort study give indications of SP from all referrers (not just GPs), providing important data to triangulate with SP data from electronic patient records. As SP programmes grow in scale internationally, the inclusion of similar measures of SP referrals in other longitudinal cohort studies is recommended.

## Data Availability

All data are publicly available through the UK Data Service

https://beta.ukdataservice.ac.uk/datacatalogue/series/series?id=200011

## Notes

### Competing Interest Statement

The authors have declared no competing interest.

### Funding Statement

This project was funded by the Nuffield Foundation Oliver Bird Fund and Versus Arthritis OBF/FR-000023818

### Author Declarations

Data are publicly available on the UK Data Service https://beta.ukdataservice.ac.uk/datacatalogue/series/series?id=200011

