## Supplementary tables for "Can social prescribing reach patients most in need? Patterns of (in)equalities in referrals in a representative cohort of older adults in England"

| Supplementary Table 1: Descriptive statistics by SP (referral); proportions and confidence intervals | | | |
| --- | --- | --- | --- |
|  | Not referred to SP | | Referred to SP |
| Age (years), mean (SE) | 65.3 (0.1) |  | 67.1 (0.5) |
| Sex |  |  |  |
| Male | 0.45 [0.44, 0.46] |  | 0.44 [0.39, 0.49] |
| Female | 0.55 [0.54, 0.56] |  | 0.56 [0.51, 0.61] |
| Marital status |  |  |  |
| Not married/cohabiting | 0.25 [0.24, 0.26] |  | 0.33 [0.28, 0.37] |
| Married/cohabiting | 0.75 [0.74, 0.76] |  | 0.67 [0.63, 0.72] |
| Working status |  |  |  |
| Not working | 0.60 [0.58, 0.61] |  | 0.68 [0.62, 0.73] |
| Working full/part-time | 0.40 [0.39, 0.42] |  | 0.32 [0.27, 0.38] |
| Receiving benefits |  |  |  |
| No | 0.86 [0.85, 0.87] |  | 0.71 [0.67, 0.76] |
| Yes | 0.14 [0.13, 0.15] |  | 0.29 [0.24, 0.33] |
| Urban dwelling |  |  |  |
| No | 0.27 [0.26, 0.28] |  | 0.21 [0.16, 0.25] |
| Yes | 0.73 [0.72, 0.74] |  | 0.79 [0.75, 0.84] |
| Index of multiple deprivation |  |  |  |
| IMD score 1-3 | 0.40 [0.39, 0.42] |  | 0.31 [0.25, 0.36] |
| IMD score 8-10 | 0.18 [0.17, 0.19] |  | 0.29 [0.24, 0.35] |
| Educational attainment |  |  |  |
| Degree | 0.28 [0.27, 0.29] |  | 0.21 [0.16, 0.26] |
| NVQ3 A level/higher education | 0.34 [0.33, 0.36] |  | 0.38 [0.33, 0.43] |
| NVQ2/GCE O level | 0.38 [0.37, 0.39] |  | 0.41 [0.35, 0.46] |
| Net non-pension wealth (tertiles) |  |  |  |
| 1 - lowest wealth quintile | 0.35 [0.33, 0.36] |  | 0.50 [0.44, 0.55] |
| 3 - highest wealth quintile | 0.32 [0.31, 0.33] |  | 0.27 [0.22, 0.32] |
| Diagnosed psychiatric condition |  |  |  |
| No | 0.92 [0.92, 0.93] |  | 0.88 [0.85, 0.91] |
| Yes | 0.08 [0.07, 0.08] |  | 0.12 [0.09, 0.15] |
| Depression (CESD>=3) |  |  |  |
| No | 0.82 [0.81, 0.83] |  | 0.69 [0.63, 0.74] |
| Yes | 0.18 [0.17, 0.19] |  | 0.31 [0.26, 0.37] |
| Diagnosed cardiovascular condition |  |  |  |
| No | 0.65 [0.64, 0.67] |  | 0.53 [0.47, 0.58] |
| Yes | 0.35 [0.33, 0.36] |  | 0.47 [0.42, 0.53] |
| Diagnosed diabetes |  |  |  |
| No | 0.92 [0.91, 0.92] |  | 0.84 [0.80, 0.88] |
| Yes | 0.08 [0.08, 0.09] |  | 0.16 [0.12, 0.20] |
| Diagnosed lung condition |  |  |  |
| No | 0.87 [0.87, 0.88] |  | 0.84 [0.80, 0.88] |
| Yes | 0.13 [0.12, 0.13] |  | 0.16 [0.12, 0.20] |
| Diagnosed MSK condition |  |  |  |
| No | 0.66 [0.65, 0.68] |  | 0.56 [0.51, 0.61] |
| Yes | 0.34 [0.32, 0.35] |  | 0.44 [0.39, 0.49] |
| Diagnosed cancer |  |  |  |
| No | 0.94 [0.93, 0.94] |  | 0.92 [0.89, 0.95] |
| Yes | 0.06 [0.06, 0.07] |  | 0.08 [0.05, 0.11] |
| Current smoker |  |  |  |
| No | 0.90 [0.89, 0.90] |  | 0.89 [0.85, 0.92] |
| Yes | 0.10 [0.10, 0.11] |  | 0.11 [0.08, 0.15] |
| Frequent/chronic pain |  |  |  |
| No | 0.61 [0.60, 0.62] |  | 0.44 [0.39, 0.49] |
| Yes | 0.39 [0.38, 0.40] |  | 0.56 [0.51, 0.61] |
| Alcohol consumption |  |  |  |
| <5 times a week | 0.77 [0.76, 0.78] |  | 0.82 [0.78, 0.86] |
| 5+ times a week | 0.23 [0.22, 0.24] |  | 0.18 [0.14, 0.22] |
| Physically inactive |  |  |  |
| No | 0.94 [0.94, 0.95] |  | 0.87 [0.83, 0.91] |
| Yes | 0.06 [0.05, 0.06] |  | 0.13 [0.09, 0.17] |
|  |  |  | . |
| *N for SP=495, N for control=6,788* | | | |

| Supplementary Table 2: Logistic Regression Model for SP (**referrals to SP**); odds ratios and confidence intervals | | | | | | | | | | | | |
| --- | --- | --- | --- | --- | --- | --- | --- | --- | --- | --- | --- | --- |
|  |  |  | 1 | | 2 | | | 3 | | | 4 | |
| Age (years) | | | 1.02 [1.01, 1.03] | *** | 1.02 [1.01, 1.04] | ** | 1.02 [1.00, 1.03] | | * | 1.02 [1.00, 1.04] | | * |
| Sex | Female | | 1.05 [0.84, 1.30] |  | 1.00 [0.81, 1.25] |  | 0.98 [0.79, 1.23] | |  | 0.99 [0.78, 1.24] | |  |
| Marital status | Married/cohabiting | |  |  | 0.87 [0.68, 1.12] |  |  | |  | 0.93 [0.72, 1.20] | |  |
| Working status | Working full/part-time | |  |  | 1.04 [0.73, 1.48] |  |  | |  | 1.14 [0.80, 1.64] | |  |
| Receiving benefits | Yes | |  |  | 2.03 [1.53, 2.70] | *** |  | |  | 1.53 [1.12, 2.08] | | ** |
| Urban dwelling | Yes | |  |  | 1.22 [0.89, 1.67] |  |  | |  | 1.21 [0.88, 1.65] | |  |
| Index of multiple deprivation | IMD score 1-3 | |  |  | 0.84 [0.61, 1.14] |  |  | |  | 0.84 [0.62, 1.15] | |  |
|  | IMD score 8-10 | |  |  | 1.39 [0.99, 1.94] |  |  | |  | 1.32 [0.94, 1.84] | |  |
| Educational attainment | NVQ3 A level/higher education | |  |  | 1.26 [0.90, 1.74] |  |  | |  | 1.22 [0.88, 1.71] | |  |
|  | NVQ2/GCE o level | |  |  | 1.02 [0.71, 1.46] |  |  | |  | 0.97 [0.67, 1.41] | |  |
| Net non-pension wealth (tertiles) | 1 - lowest wealth quintile | |  |  | 1.55 [1.13, 2.13] | ** |  | |  | 1.43 [1.03, 1.99] | | * |
|  | 3 - highest wealth quintile | |  |  | 1.33 [0.94, 1.86] |  |  | |  | 1.39 [0.99, 1.96] | |  |
| Diagnosed psychiatric condition | Yes | |  |  |  |  | 1.20 [0.85, 1.71] | |  | 1.16 [0.81, 1.65] | |  |
| Depression (CESD>=3) | Yes | |  |  |  |  | 1.60 [1.20, 2.12] | | ** | 1.44 [1.07, 1.94] | | * |
| Diagnosed cardiovascular condition | Yes | |  |  |  |  | 1.27 [0.99, 1.61] | |  | 1.23 [0.96, 1.56] | |  |
| Diagnosed diabetes | Yes | |  |  |  |  | 1.52 [1.11, 2.10] | | * | 1.39 [1.00, 1.94] | | * |
| Diagnosed lung condition | Yes | |  |  |  |  | 1.01 [0.73, 1.41] | |  | 0.97 [0.70, 1.36] | |  |
| Diagnosed MSK condition | Yes | |  |  |  |  | 1.04 [0.82, 1.33] | |  | 1.00 [0.78, 1.28] | |  |
| Diagnosed cancer | Yes | |  |  |  |  | 1.08 [0.71, 1.66] | |  | 1.07 [0.69, 1.65] | |  |
| Current smoker | Yes | |  |  |  |  | 1.04 [0.72, 1.50] | |  | 0.88 [0.60, 1.30] | |  |
| Frequent/chronic pain | Yes | |  |  |  |  | 1.57 [1.22, 2.02] | | *** | 1.45 [1.12, 1.88] | | ** |
| Alcohol consumption | 5+ times a week | |  |  |  |  | 0.81 [0.62, 1.04] | |  | 0.82 [0.63, 1.07] | |  |
| Physically inactive | Yes | |  |  |  |  | 1.63 [1.10, 2.41] | | * | 1.34 [0.89, 2.02] | |  |
| Intercept | | | 0.02 [0.01, 0.04] | *** | 0.01 [0.00, 0.03] | *** | 0.02 [0.01, 0.04] | | *** | 0.01 [0.00, 0.03] | | *** |
| Number of observations | | | 7283 |  | 7283 |  | 7283 | |  | 7283 | |  |
| **** p<.001, ** p<.01, * p<.05 Column 1: adjusted for age and sex Column 2: adjusted for model 1 + socio-demographic factors Column 3: adjusted for model 1 + health factors Column 4: adjusted for all factors Reference categories: Educational attainment - degree; wealth - mid quintile* | | | | | | | | | | | | |

| Supplementary Table 3: Logistic Regression Model for SP (**uptake of SP**); odds ratios and confidence intervals | | | | | | | | | | |
| --- | --- | --- | --- | --- | --- | --- | --- | --- | --- | --- |
|  |  |  | 1 | | 2 | | 3 | | 4 | |
| Age (years) | | | 1.02 [1.01, 1.04] | *** | 1.02 [1.01, 1.04] | ** | 1.02 [1.00, 1.03] | * | 1.02 [1.00, 1.04] | * |
| Sex | Female | | 1.05 [0.83, 1.32] |  | 1.00 [0.79, 1.27] |  | 1.00 [0.78, 1.27] |  | 0.99 [0.77, 1.27] |  |
| Marital status | Married/cohabiting | |  |  | 0.85 [0.65, 1.10] |  |  |  | 0.89 [0.68, 1.16] |  |
| Working status | Working full/part-time | |  |  | 0.95 [0.69, 1.33] |  |  |  | 1.03 [0.74, 1.43] |  |
| Receiving benefits | Yes | |  |  | 1.98 [1.47, 2.67] | *** |  |  | 1.55 [1.12, 2.14] | ** |
| Urban dwelling | Yes | |  |  | 1.14 [0.81, 1.60] |  |  |  | 1.13 [0.81, 1.59] |  |
| Index of multiple deprivation | IMD score 1-3 | |  |  | 0.78 [0.57, 1.07] |  |  |  | 0.78 [0.57, 1.07] |  |
|  | IMD score 8-10 | |  |  | 1.39 [0.96, 2.01] |  |  |  | 1.31 [0.90, 1.91] |  |
| Educational attainment | NVQ3 A level/higher education | |  |  | 1.23 [0.88, 1.72] |  |  |  | 1.20 [0.85, 1.68] |  |
|  | NVQ2/GCE o level | |  |  | 1.00 [0.69, 1.45] |  |  |  | 0.96 [0.65, 1.40] |  |
| Net non-pension wealth (tertiles) | 1 - lowest wealth quintile | |  |  | 1.43 [0.99, 2.05] |  |  |  | 1.33 [0.92, 1.92] |  |
|  | 3 - highest wealth quintile | |  |  | 1.36 [0.93, 2.00] |  |  |  | 1.43 [0.97, 2.11] |  |
| Diagnosed psychiatric condition | Yes | |  |  |  |  | 1.13 [0.76, 1.68] |  | 1.07 [0.72, 1.60] |  |
| Depression (CESD>=3) | Yes | |  |  |  |  | 1.45 [1.08, 1.94] | * | 1.30 [0.96, 1.77] |  |
| Diagnosed cardiovascular condition | Yes | |  |  |  |  | 1.23 [0.95, 1.60] |  | 1.20 [0.92, 1.55] |  |
| Diagnosed diabetes | Yes | |  |  |  |  | 1.62 [1.15, 2.27] | ** | 1.49 [1.05, 2.10] | * |
| Diagnosed lung condition | Yes | |  |  |  |  | 1.02 [0.73, 1.43] |  | 0.98 [0.70, 1.38] |  |
| Diagnosed MSK condition | Yes | |  |  |  |  | 1.03 [0.80, 1.33] |  | 0.98 [0.76, 1.27] |  |
| Diagnosed cancer | Yes | |  |  |  |  | 1.08 [0.70, 1.68] |  | 1.06 [0.67, 1.66] |  |
| Current smoker | Yes | |  |  |  |  | 1.12 [0.75, 1.69] |  | 0.96 [0.62, 1.48] |  |
| Frequent/chronic pain | Yes | |  |  |  |  | 1.58 [1.21, 2.06] | *** | 1.48 [1.12, 1.94] | ** |
| Alcohol consumption | 5+ times a week | |  |  |  |  | 0.83 [0.63, 1.08] |  | 0.85 [0.65, 1.11] |  |
| Physically inactive | Yes | |  |  |  |  | 1.46 [0.96, 2.23] |  | 1.20 [0.78, 1.84] |  |
| Intercept | | | 0.01 [0.01, 0.03] | *** | 0.01 [0.00, 0.03] | *** | 0.01 [0.00, 0.03] | *** | 0.01 [0.00, 0.03] | *** |
| Number of observations | | | 7283 |  | 7283 |  | 7283 |  | 7283 |  |
| **** p<.001, ** p<.01, * p<.05 Column 1: adjusted for age and sex Column 2: adjusted for model 1 + socio-demographic factors Column 3: adjusted for model 1 + health factors Column 4: adjusted for all factors Reference categories: Educational attainment - degree; wealth - mid quintile* | | | | | | | | | | |
